# Clonal hematopoiesis is associated with distinct rheumatoid arthritis phenotypes

**DOI:** 10.1101/2024.10.10.24315184

**Authors:** E Hiitola, J Korhonen, H Kokkonen, J Koskela, M Kankainen, M Alakuijala, A Liu, S Lundgren, P Häppölä, H Almusa, P Ellonen, P Savola, T Kelkka, M Leirisalo-Repo, R Koivuniemi, R Peltomaa, L Pirilä, P Isomäki, M Kauppi, O Kaipiainen-Seppänen, I Starskaia, AT Virtanen, R Lahesmaa, O Silvennoinen, FinnGen, G Genovese, A Ganna, S Rantapää-Dahlqvist, S Mustjoki, M Myllymäki

## Abstract

Clonal hematopoiesis (CH) becomes more prevalent with age and may impact the pathophysiology of inflammatory diseases by altering immune cell function. While clonal hematopoiesis of indeterminate potential (CHIP) can promote inflammation in non-malignant conditions, the relationship with rheumatoid arthritis (RA) has not been systematically investigated. We tested whether CHIP mutations are more common in RA using two population-level cohorts and newly diagnosed RA patients. CHIP was associated with prevalent RA in the FINRISK study of 10 089 participants with whole exome sequencing (odds ratio (OR)=2.06, 95% CI=1.08-3.94, P=0.029) and in the FinnGen cohort (N=520 210, OR=1.42, 95% CI=1.09-1.84, P=0.009) using single nucleotide polymorphism (SNP) array-based CHIP annotation. In the FinnGen cohort, *DNMT3A* mutations were associated with seropositive RA (OR=1.73, 95% CI=1.16-2.58, P=0.007), whereas CHIP overall was more common in participants with history of seronegative RA (OR=2.16, 95% CI=1.24-3.76, P=0.006). Furthermore, CHIP was associated with inferior overall survival among FinnGen participants with prevalent RA (P=0.010). In newly diagnosed RA (N=632), seropositive, but not seronegative, patients with *DNMT3A* mutations had higher erythrocyte sedimentation rate (P=0.014) and disease-activity scores (P=0.030). In contrast, *TET2* mutations were significantly more common in patients with seronegative RA both in univariable (P=0.009) and multivariable models (OR=0.42; 95% CI=0.20-0.89, P=0.024). In conclusion, CHIP is associated with RA and distinct RA subtypes. Although the causality and underlying mechanisms of these observations remain unknown, our findings provide further evidence for the association between CHIP and inflammation in distinct disease contexts that may have therapeutic implications in the future.

## Introduction

The role of aging-related clonal hematopoiesis (CH) in inflammatory diseases remains to be fully characterized^1,2^. Clonal hematopoiesis of indeterminate potential (CHIP) is defined by the presence of hematopoietic stem cell (HSC) clones carrying somatic mutations in genes recurrently mutated in myeloid neoplasms^3^. CHIP predisposes to hematologic malignancies^3^, but the increased risk of death among CHIP carriers^4^ is mainly caused by non-malignant inflammatory phenotypes, such as cardiovascular diseases (CVD)^5^. CHIP can promote inflammation via multiple mechanisms, such as via hyperactivation of the NLR family pyrin domain containing 3 (NLRP3) inflammasome and subsequent cytokine release from monocytes/macrophages^6^. Consequently, CHIP has been implicated in the pathogenesis of inflammatory diseases such as gout^7^, chronic liver disease^8^, acute kidney injury^9^, giant cell arteritis^10^, and CVD^4,11^. On the other hand, chronic inflammation can boost the expansion of CHIP clones in the bone marrow^12,13^. Anti-inflammatory therapies have shown preliminary efficacy in CVD prevention in high-risk patients with CHIP^14^, suggesting that identifying CHIP carriers may result in personalized therapeutic opportunities in inflammation-driven disease contexts. In addition to CHIP, other CH subtypes include mosaic chromosomal alterations (mCAs)^15,16^ and CH characterized by somatic variants in lymphoid driver genes^17,18^. Both mCAs^19^ and lymphoid driver variants^17^ increase the risk of hematological cancers, but their impact on other disease phenotypes remains to be explored.

Rheumatoid arthritis (RA), one of the most common autoimmune disease primarily affecting joints^20^, significantly co-occurs with chronic myeloid malignancies^21,22^. Seropositive RA is defined by the presence of anti-cyclic citrullinated peptide antibodies (ACPA) and/or rheumatoid factor (RF), whereas seronegative RA is a more heterogeneous inflammatory disease entity involving patients with clinical RA manifestations in the absence of ACPA and RF^23^. Clonal expansions of the immune cells have been reported in RA. For example, patients with Felty syndrome, a disease entity characterized by neutropenia, splenomegaly, and seropositive RA, have a high prevalence of *STAT3* mutations in peripheral blood^24^. CHIP has also been associated with autoimmune diseases in 200 patients who underwent hip arthroplasty for osteoarthritis^25^, and with RA in a study of 1794 participants aged 80 or older^26^. However, these preliminary observations have yet to be validated in population-level cohorts, and studies on the prevalence of CHIP in clinically annotated RA patients has so far been limited to a small patient cohort of 59 RA patients^27^. Understanding the association between CHIP and RA may provide novel insights into the pathophysiology, prevention, and clinical management of autoimmune diseases.

In this study, we comprehensively analyzed whether CH subtypes are associated with RA in the population level using whole-exome sequencing data in the FINRISK study^28^ and SNP array data in the FinnGen cohort^29^. We also performed targeted sequencing of CHIP genes in a cohort of 632 previously untreated RA patients and 163 healthy controls to elucidate the prevalence of CHIP in distinct RA phenotypes (Supplementary Table 1). Collectively, our approaches highlight the spectrum of CH associated with RA in a context-dependent manner.

## Methods

### FINRISK cohort

The FINRISK study is a Finnish population-level cohort consisting of random population-level sampling of individuals aged 25-74^28^. We analyzed disease endpoints from Finnish national health registries, laboratory measurements, and questionnaire-based information for the FINRISK participants recruited between 1992 and 2007.

### Somatic variant calling of CHIP and lymphoid driver variants in FINRISK

We obtained whole exome sequencing (WES) data for 10 129 previously sequenced FINRISK participants^30^. We called CH variants from the WES data using Illumina DRAGEN v3.8 in tumor-only mode. We generated a panel-of-normals using 40 FINRISK participants under 40 years of age, and these samples were confirmed not to have any putative CHIP variants. The panel of normals was then included in the Illumina Dragen variant calling algorithm. We used Annovar for variant annotation. Variant calling, annotation and filtering parameters are described in detail in the Supplementary Methods.

### FinnGen cohort

FinnGen is a research project^29^ that has compiled genotype and phenotype information from 520 210 Finnish individuals (FinnGen Data Freeze 12). The sample collection and genotyping performed in the FinnGen study are described in the Supplementary Methods. The genetic information available is combined with data collected from Finnish national health registries, including diagnosis codes.

### CHIP calling in FinnGen

CHIP hotspot variants were called in FinnGen by manually examining SNP intensity plots in coding areas in CHIP associated genes. We analyzed candidate loci in CHIP genes covered by the FinnGen SNP array. Loci were further evaluated by calculating associations with age, hematologic malignancy, and the relative prevalence to detected variants in the publicly available UK Biobank (UKBB) WES data^31^ (Supplementary Methods and Supplementary Table 8). The relative prevalences of the variants were also compared with CHIP in the TOPMED whole genome sequencing data^32^.

### mCA detection in FINRISK and FinnGen

mCAs were called in FINRISK and FinnGen using the MoChA pipeline^15^ with SHAPEIT4^33^ haplotype phasing. Samples with a call rate of less than 0.97 or BAF autocorrelation of more than 0.03 were excluded.

### Endpoint definitions in FINRISK and FinnGen

Disease endpoints were defined by querying national health registries for ICD-codes. ICD-codes defining endpoints are defined in Supplementary Table 6. RA endpoints in FINRISK and FinnGen were annotated as seronegative only if there were no codes for seropositive RA in the registries, i.e. participants with both codes found in the registry were defined as seropositive.

### RA patient cohorts

We compiled a cohort of 573 newly diagnosed RA patients with peripheral blood sample available at RA diagnosis from the Rheumatology Biobank of Umeå, Sweden, the Finnish FIN-RACo^34^ and NEO-RACo^35^ clinical trials, and a prospective cohort of 45 Finnish participants (FosfoRA study). Together with the cohort from Savola et al^27^ we analyzed a total of 632 RA patients. The median age of the RA patients was 64 years (18-89) and the median year of diagnosis was 2006 (1993-2021). 65% of RA patients were female, and 434 out of 632 RA patients were annotated as seropositive. Characteristics of the individual cohorts are listed in the Supplementary Methods and in Supplementary Table 9.

### Targeted next-generation sequencing panel to detect CHIP

We identified CHIP variants in RA patients and healthy controls from the Finnish Red Cross Blood Service Biobank using panel sequencing of 65 myeloid driver genes (Supplementary Table 10). Illumina NovaSeq 6000 system was used for sequencing, and the median target sequencing coverage was 1700x across samples. Further details of the DNA processing, sequencing and variant calling is described in the Supplementary Methods.

### Statistical methods

In each sub cohort, genotypes were assigned before evaluating the association with clinical variables, including outcomes. Analyses were performed using Python and GraphpadPrism. For FinnGen data, analyses were performed in the dedicated Sandbox environment. Python package statsmodels 0.14.2 was used for fitting logistic regression models, lifelines 0.29.0 was used for fitting Cox proportional hazards (Cox-PH) and Kaplan Meier models, and statistical tests implemented in scipy 1.11.4 and GraphPad Prism 10.3.0 were used. P-values reported are 2-sided. P values less than 0.05 were considered statistically significant throughout the study unless otherwise mentioned. Multivariable models were adjusted for age, sex, smoking and 10 principal components of ancestry, and participants with missing values in any of these were excluded, unless otherwise mentioned. Hematologic malignancies were excluded in co-occurrence models and right censored in survival models.

### Data availability

Access to FinnGen and FINRISK data can be requested by contacting the FinnGen consortium and the Finnish Institute for Health and Welfare (THL) Biobank, respectively. The data for RA patient cohorts can be requested from the respective biobanks. According to constraints in the ethical permit, sequencing data of patients is only available from the corresponding author upon reasonable request.

## Results

### Clonal hematopoiesis in the FINRISK cohort

To evaluate the association between CH and RA, we reanalyzed WES^30^ and SNP array data to detect CHIP and mCAs, respectively, in the FINRISK cohort of 10 129 participants that are highly representative of the Finnish population^30^ (Methods). The median age of FINRISK participants with WES available was 49 years (range, 25 to 74) and 53 percent of participants were male (Supplementary Figure 1A). The median registry follow-up time in FINRISK participants was 15.8 years after DNA sampling (range 0-25.9 years).

Among the 10 089 FINRISK participants with WES data and not in the panel of normals, the median exome target coverage was 48x (Supplementary Figure 1C). We identified a total of 411 variants in CHIP genes in 381 FINRISK participants (Supplementary Tables 2 and 4, Supplementary Figure 2). Among those with CHIP, most participants (352/381, 92%) had one CHIP variant (Supplementary Figure 2G). Consistent with prior reports^31,32^, the most recurrently mutated genes included *DNMT3A*, *TET2*, and *ASXL1* (Figure 1A). The ratio of *DNMT3A* variants to *TET2* variants was lower in our cohort compared with UKBB^31^, but the difference was less evident when adjusting for coverage of the target regions (Supplementary Figure 2H). Most mutations were missense (Supplementary Figure 2A), and the most common nucleotide substitution was cytosine to thymine (C>T) (Supplementary Figure 2B). The association between CHIP variants and sex was also consistent with the UKBB data^31^ (Supplementary Figure 2E). The distribution of variant allele frequencies (VAFs) by mutated genes and variant consequences are listed in Supplementary Figure 2C-D. Notably, we observed no somatic hotspot variants in *UBA1*, a gene underlying VEXAS syndrome characterized by autoimmune phenotypes^38,39^ as expected based on recently reported population prevalence of *UBA1* mutations in the United States^40^.

**Figure 1:**
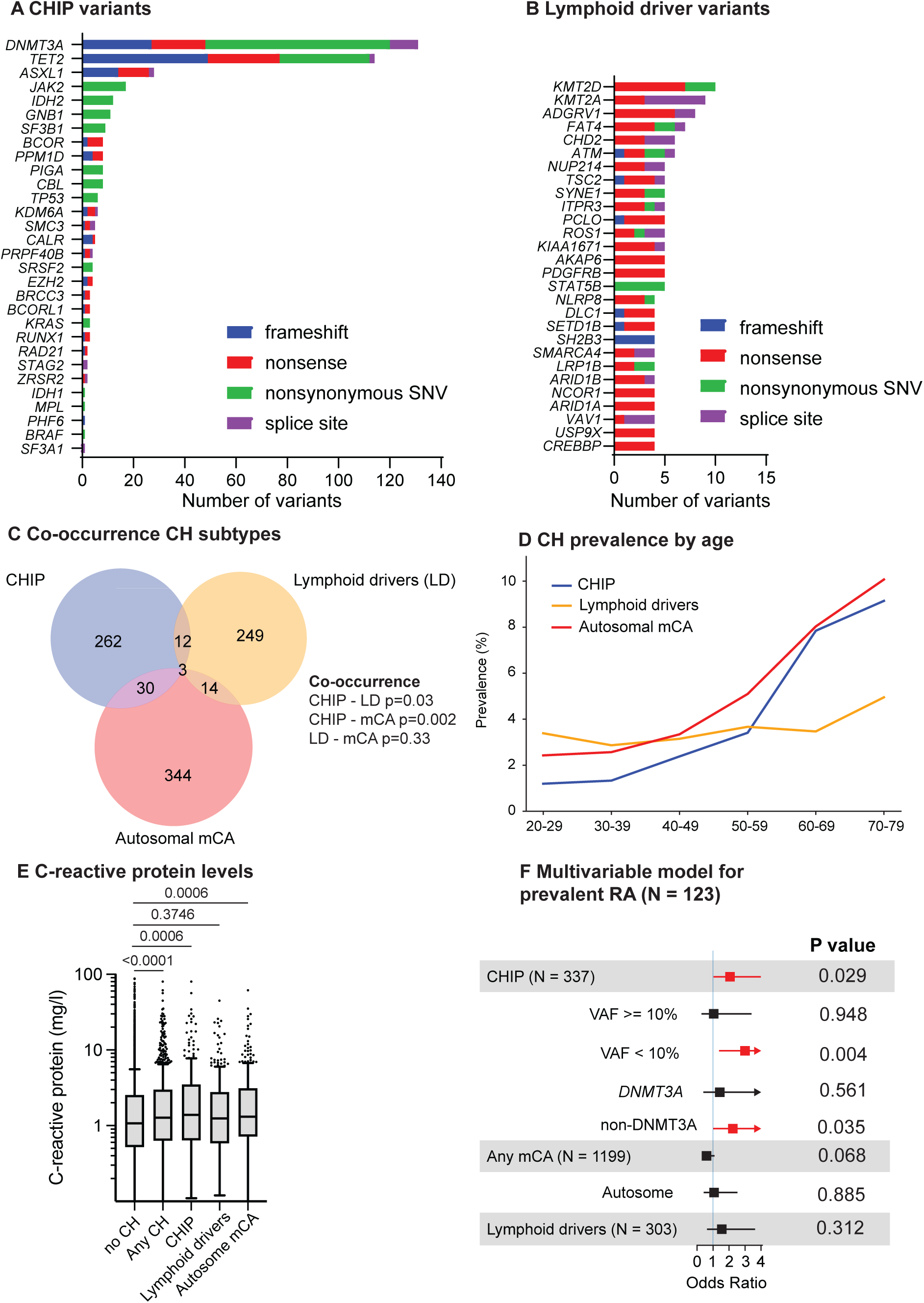
Spectrum of clonal hematopoiesis and association with RA in the FINRISK study. (A) Distribution of CHIP variants by gene and variant type in the FINRISK study. (B) Number of variants by variant type in putative lymphoid driver genes in FINRISK. (C) Co-occurrence of CH subtypes. P values were calculated using Fisher’s exact test. (D) Prevalence of CH subtypes in FINRISK participants by age in decades. (E) C-reactive protein levels by CH subtypes. (F) Associations between CH subtypes and prevalent RA diagnosis, adjusted for age, sex, smoking and 10 principal components of genetic ancestry. Participants with prevalent hematological cancers were excluded from the analyses of panels C to F. See Supplementary figures 2 and 3 for additional characteristics of CHIP and lymphoid driver variants, respectively. CH: Clonal hematopoiesis, CHIP: clonal hematopoiesis of indeterminate potential, mCA: mosaic chromosomal alterations, VAF: Variant allele frequency.

We also evaluated the spectrum of lymphoid driver variants^17,41^ in the FINRISK cohort (Methods, Supplementary Table 3, Figure 1B). We identified 354 variants in 346 participants in lymphoid driver genes (Supplementary Table 4, Supplementary Figure 3). The most mutated lymphoid driver genes included *KMT2D*, *KMT2A*, and *ADGRV1*; however, only a total of 27 variants were observed in these genes (Figure 1B). Most mutations were nonsense (Supplementary Figure 3A), and the most prevalent nucleotide substitution was C>A (Supplementary Figure 3B). The distribution of VAFs by mutated lymphoid driver genes and variant consequences are listed in Supplementary Figure 3C-D. Most of the participants with lymphoid driver variants had only one variant (Supplementary Figure 3E). The median VAFs were lower for putative lymphoid driver versus myeloid CHIP variants (6.8% vs 9.1%, P value <0.001, Supplementary Figure 3F), consistent with lymphoid driver variants likely occurring in lymphoid progenitor/differentiated cells versus CHIP variants occurring at stem/progenitor cell level.

In addition, we called mCAs in 8414 FINRISK participants with available SNP array genotyping data (Methods) and found 406 autosomal mCAs in 402 participants (Supplementary Table 5, Supplementary Figure 4). We identified loss of the sex chromosomes as the most common mCAs and CN-LOH in 1q, 2q and 14q as the most common autosomal mCAs (Supplementary Figure 4). CHIP significantly co-occurred with lymphoid drivers (OR=1.69, P=0.03) as well as with autosomal mCAs (OR=1.86, P=0.002) in multivariable models. Lymphoid drivers did not significantly co-occur with autosomal mCAs (OR=1.29, P=0.33) (Figure 1C).

### Clonal hematopoiesis and clinical parameters in FINRISK

The prevalence of CHIP was strongly age-related, whereas the prevalence of lymphoid drivers showed no evident age-association (Figure 1D). CHIP prevalence reached 7.8 percent (95% CI=6.6-9.1%) in participants aged 60-69, consistent with the previously reported prevalence in population cohorts^31^.

We systematically evaluated the association between CH and laboratory variables measured as part of FINRISK (Supplementary Figure 5A). Participants with CHIP or autosomal mCAs had higher C-reactive protein levels in univariable models (Figure 1E), in line with previous findings^42^, suggesting that CHIP may be linked to inflammatory processes.

As expected, myeloid and lymphoid driver variants were associated with prevalent and incident hematologic malignancies (Supplementary Figure 5C-D). In addition, CHIP mutations were associated with heavy but not mild smoking history, that was mostly driven by mutations in *ASXL1,* as previously reported^43,44^. Autosomal mCAs were also associated with heavy smoking history^45^ (Supplementary Figure 5B).

### CHIP is enriched in FINRISK participants with RA

After excluding participants with prevalent hematologic malignancies, 1.2% (121/10021) FINRISK participants had prevalent RA at DNA sampling. The presence of CHIP mutations was significantly associated with prevalent RA (OR=2.06, P=0.029; 95% CI=1.08-3.94) in a multivariable model (Figure 1F). This association was specific to participants with maximum VAF of CHIP variants less than 10% (OR=3.00, P=0.004, 95% CI, 1.41-6.35) and was driven by non-*DNMT3A* CHIP mutations (Figure 1F). The association was validated in questionnaire-based annotation of prevalent RA (Supplementary Figure 6A). In contrast, the presence of lymphoid driver variants or mCA subtypes were not associated with prevalent RA (Figure 1F). Nine patients with CHIP developed incident RA during follow-up (HR=1.62, 95% CI 0.82-3.21, P=0.16) (Supplementary Figure 6B). Taken together, CHIP is more common in participants with prevalent RA in the FINRISK cohort.

### Deriving CHIP status in SNP arrays

DNA SNP arrays have been used to derive CHIP carriers based on outlier status in intensity plots at loci covered by the array^8^. To systematically evaluate the concordance between array-derived CHIP status and CHIP detection with next-generation sequencing, we analyzed *JAK2* V617F and *DNMT3A* R882H, the two most recurrent CHIP hotspot loci^31^, in SNP intensity and WES data in the FINRISK cohort. 8 out of 11 (74%) CHIP carriers in *JAK2* V617F, and 12 out of 29 (41%) CHIP carriers in *DNMT3A* R882H based on WES were also independently captured by detecting outliers in the SNP intensity plots (Figure 2A, Supplementary Table 7). Variants at *JAK2* V617F and *DNMT3A* R882H with a VAF of 10% or more were detected with 77% and 90% likelihood in the SNP array data, respectively (Figure 2A, Supplementary Table 7). Among participants with CHIP in FINRISK WES data, SNP array - derived relative B allele frequencies showed a strong correlation with WES-derived VAFs in both CHIP hotspots analyzed (Figure 2B). Based on these analyses in the FINRISK cohort, we estimate that a high proportion (80-90% with VAF > 10%) of CHIP hotspot variants covered in the SNP array can be detected by manual curation of SNP intensity plots in a clone size-dependent manner.

**Figure 2:**
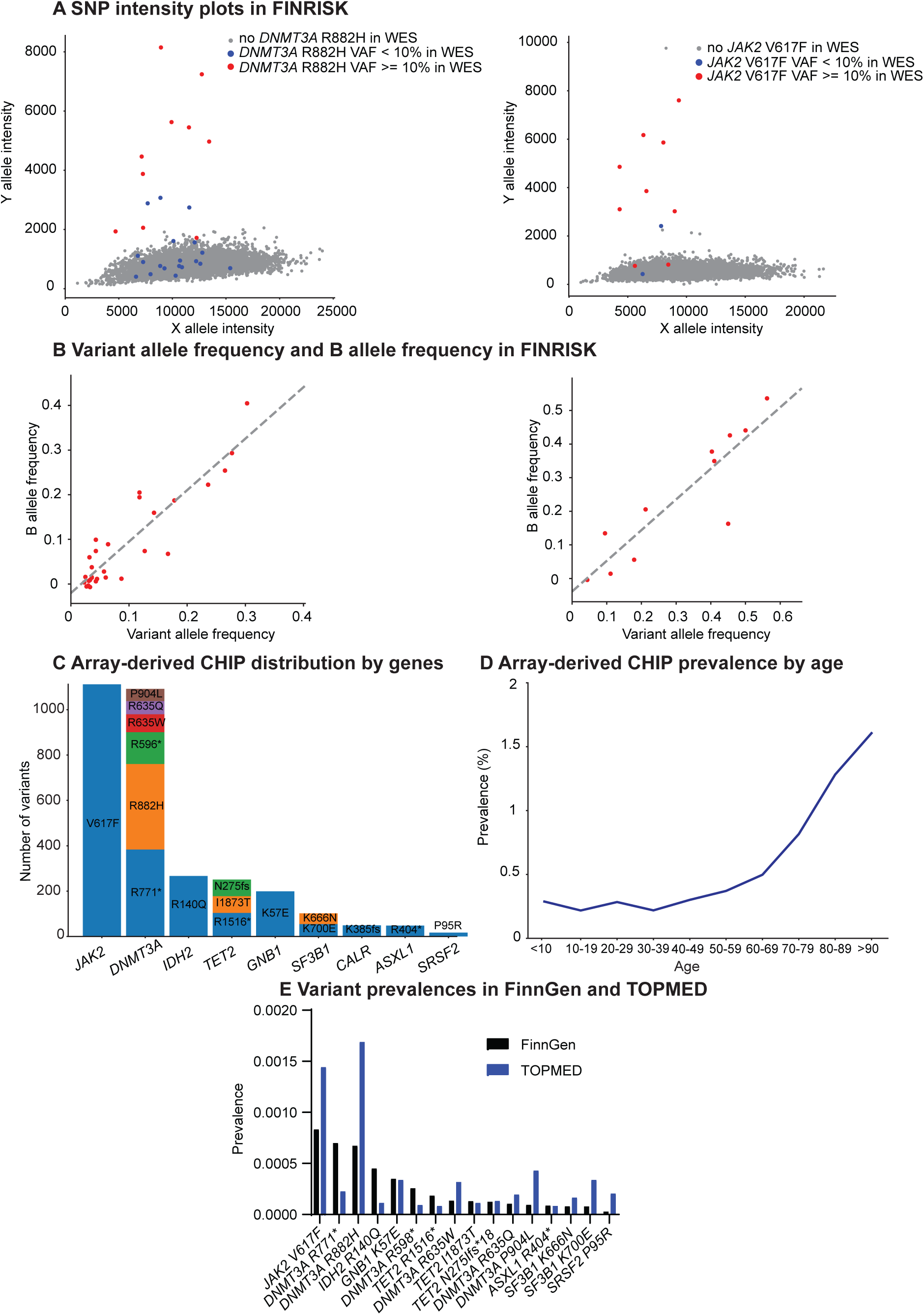
Detection of array-derived CHIP in FinnGen. (A) SNP intensity plots of CHIP hotspot variants *DNMT3A* R882H and *JAK2* V617F in FINRISK. Blue dots represent participants with the variant detected in WES with VAF < 10%, red dots represent VAF >= 10%, and grey dots represent no detected variant in WES. (B) Correlation of SNP array-derived B allele frequencies and WES-derived variant allele frequencies at *DNMT3A* R882H and *JAK2* V617F in FINRISK. P-values calculated using Wald test. (C) Distribution of array-derived CHIP hotspot variants in FinnGen. (D) Array-derived CHIP prevalence by age in FinnGen, excluding prevalent hematologic malignancies. (E) Comparison of detected hotspot variant prevalences between FinnGen and TOPMED, excluding prevalent hematologic malignancies. SNP: Single nucleotide polymorphism, WES: Whole-exome sequencing.

The FinnGen cohort consists of SNP array data and registry-level clinical information for over 500 000 Finnish participants, covering almost ten percent of the Finnish population^29^. The relatively old participant age (median, 55 years) and high proportion of hospital-based recruitment facilitates enrichment of prevalent disease endpoints in FinnGen^29^. A total of 49 loci in the coding regions of CHIP genes^31^ were covered by the FinnGen SNP array. We manually inspected intensity plots in all these sites, and used heuristics, including variant prevalence relative to UKBB data^31^ and associations with age and hematologic malignancies, to identify 17 high-confidence CHIP loci (Methods, Supplementary Methods, Supplementary Table 8, Figure 2C). As expected, the *JAK2* V617F had the highest CHIP hotspot prevalence, followed by *DNMT3A* R771* and R882H (Figure 2C). The prevalence of FinnGen participants with array-derived CHIP reached 0.8 percent by age 70 years (Figure 2D, Supplementary Figure 7B). CHIP carrier status was associated with prevalent and incident hematologic malignancies (Supplementary Figure 7C-D), but not with higher CRP levels (Supplementary Figure 7E). As an additional control, we confirmed similar prevalence of variants in our high-confidence loci with previously published whole-genome sequencing data that robustly detected CHIP variants with >10% VAF from the TOPMED cohort^32^ (Figure 2E). The median age of participants was 55 years in both FinnGen and TOPMED cohorts. Taken together, detecting CHIP hotspot variants in FinnGen SNP array data is feasible but has limited sensitivity for smaller clones.

### CHIP is more common in FinnGen participants with seronegative RA

12636 (2.5%) FinnGen participants had a history of RA diagnosis prior to DNA sampling (Supplementary Figure 8A). Consistent with results in the FINRISK cohort, CHIP carrier status was associated with prevalent RA in a multivariable model in FinnGen (OR=1.42, P=0.009, 95% CI,1.09-1.84) (Figure 3A). This association was specific to variants in *DNMT3A*, but not in *TET2* or *JAK2* (Supplementary Figure 8B). We observed no signal between autosomal mCAs and prevalent RA (Fig 3A, Supplementary Figure 9). With 3655 incident RA cases, we found no difference in the cumulative incidence of RA by CH status in the FinnGen cohort (Supplementary Figure 10).

**Figure 3:**
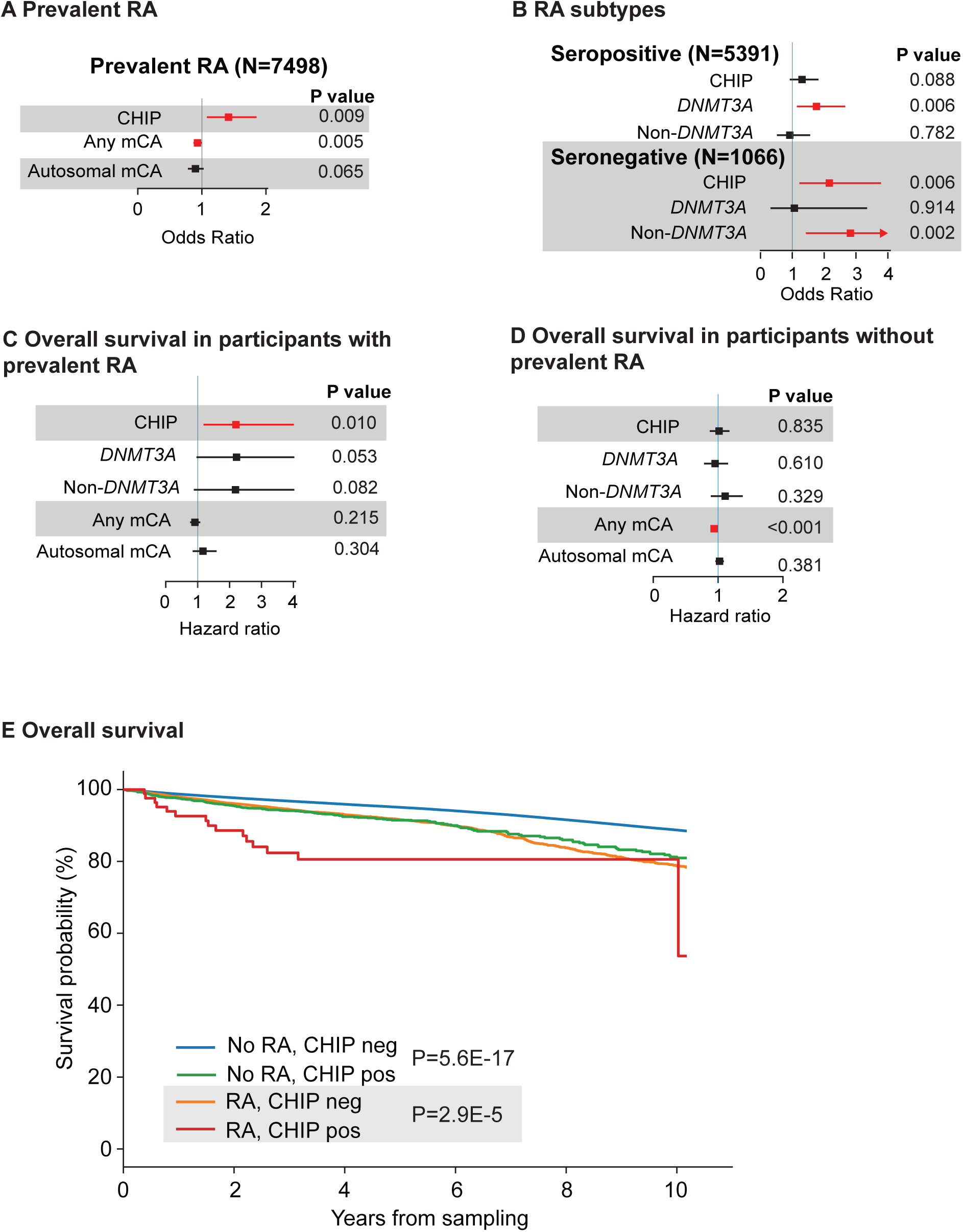
Association between CHIP and RA in FinnGen. (A) Association between CHIP and RA diagnosis prior to sampling. (B) Association between CHIP and RA subtype diagnosis prior to sampling. (C) Cox-PH model for overall mortality in FinnGen participants with prevalent RA diagnosis. (D) Cox-PH model for overall mortality in FinnGen participants without prevalent RA diagnosis. A-D adjusted for age, sex, smoking, principal components for ancestry and excluded or censored for hematologic malignancies. In addition, the multivariable model in C was adjusted for time from RA diagnosis to sampling. (E) Kaplan Meier of overall survival in FinnGen participants. C-E Outcomes calculated from time of sample collection

Interestingly, array-derived CHIP status was associated with prevalent seronegative (N of prevalent seronegative cases, 1066; OR=2.16, 95% CI=1.24-3.76, P=0.006) but not with seropositive (N of prevalent seropositive cases, 5391; OR=1.29, 95% CI=0.95-1.77, P=0.11) RA (Figure 3B). CHIP carrier status in *DNMT3A* was specifically associated with seropositive RA (OR=1.73, 95% CI=1.16-2.58, P=0.007); whereas non-DNMT3A CHIP was associated with seronegative RA (OR=2.82, 95% CI=1.45-5.48, P=0.002) in multivariable models (Figure 3B).

### CHIP confers worse survival among RA patients

Among FinnGen participants with prevalent RA, the presence of CHIP conferred inferior OS when evaluated from the time of DNA sampling in a multivariable model (Figure 3C). Interestingly, array-derived CHIP was not associated with OS in FinnGen participants without RA (Figure 3D), suggesting that CHIP may increase the mortality rate specifically in RA patients. The 5-year OS from DNA sampling for RA patients was 81% in CHIP carriers versus 92% in non-CHIP carriers (Figure 3E).

### CHIP in newly diagnosed RA

To further evaluate the clinical impact of CHIP on RA phenotypes and outcomes, we collected DNA samples from four distinct RA patient cohorts of newly diagnosed, previously untreated, RA patients without hematologic malignancies (Supplementary Table 9). A total of 573 RA patients and 163 healthy controls underwent targeted NGS sequencing using a sequencing panel consisting of 65 genes recurrently mutated in myeloid malignancies (Supplementary Tables 9-10). We also included a previously published cohort of 59 RA patients with CHIP information^27^. The median age of RA patients included was 63 years.

We identified total of 204 CHIP variants in 134 RA patients; 99 of those patients only had one variant (Supplementary Figure 11B, Supplementary Table 11). The overall prevalence of CHIP variants in newly diagnosed RA patients was 22% (126/573, Figure 4A). As expected^5^, the most commonly mutated genes were *DNMT3A*, *TET2*, *ASXL1* (Figure 4B). We observed that the highest median VAFs in *ASXL1*, S*RSF2*, *SF3B1*, and *GNAS* (Figure 4C). We observed no differences in CHIP prevalence between RA patients and healthy controls (Supplementary Figure 11C).

**Figure 4:**
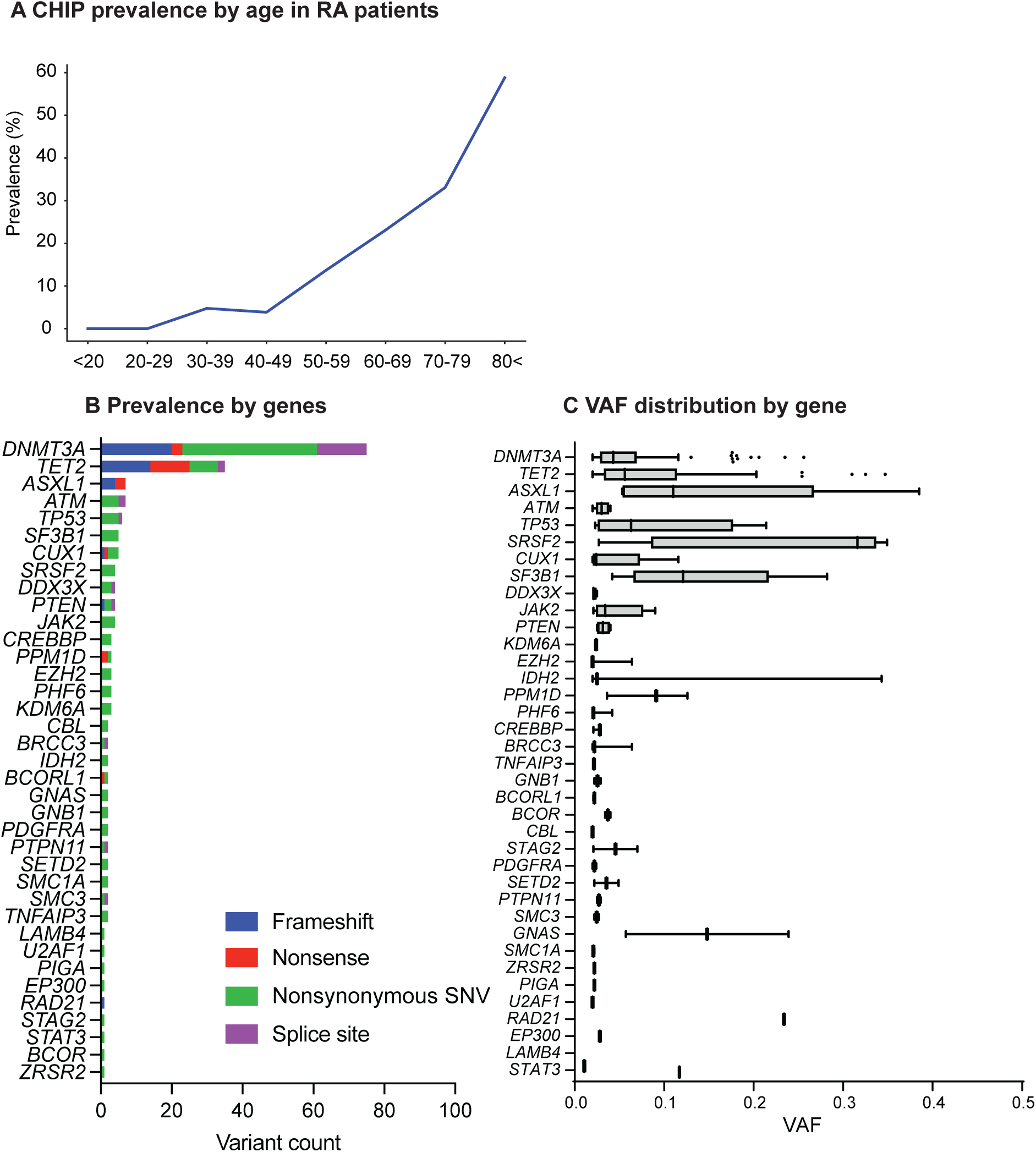
Characterising CHIP in RA patients. (A) CHIP prevalence by age in RA patient cohort (N = 632). (B) Distribution of CHIP variants by gene and variant type. (C) Tukey box plot of variant allele frequencies by mutated gene in RA patient cohort. Cohort characteristics are listed in Supplementary Table 9.

Next, we evaluated RA patient characteristics by CHIP status. The median age of RA patients with CHIP was 69 years, versus 62 years in RA patients without CHIP (P<0.001) (Table 1). While we observed no differences in C-reactive protein or erythrocyte sedimentation rate (ESR) (Table 1), the level of functional ability was lower, as indicated by higher health assessment questionnaire (HAQ) scores in RA patients with CHIP versus no CHIP (P=0.01) (Table 1). The association with higher HAQ scores remained significant when adjusting for age and sex (P=0.009). We observed no difference in the prevalence of comorbidities by CHIP status among RA patients (Table 1). Among 15 participants with coronary artery CT scan available as part of the ERA CVD study, we observed no difference by CHIP status in the coronary artery score (P=0.44) (Supplementary Figure 12).

**Table 1.** Patient characteristics of newly diagnosed RA patients.

|  | **N patients with information available** | **No CHIP (N=447)** | **CHIP (N=126)** | **P value** |
| --- | --- | --- | --- | --- |
| **Patient characteristics** |  |  |  |  |
| Age at diagnosis, median (range) | 573 | 62 (18-86) | 69 (38-89) | 5.74E-10 |
| Male sex, n (%) | 573 | 162 (36) | 47 (37) | 0.83 |
| Smoking history, n (%) | 487 | 221 (60) | 60 (51) | 0.11 |
| BMI, median (range) | 489 | 26.1 (17.2-44.8) | 26.1 (17.0-37.2) | 0.98 |
| **RA characteristics** |  |  |  |  |
| RF+, n (%) | 493 | 235 (63) | 69 (58) | 0.39 |
| ACPA+, n (%) | 495 | 229 (61) | 65 (55) | 0.24 |
| Seropositive, n (%) | 555 | 302 (70) | 80 (64) | 0.19 |
| Baseline CRP, median (range) | 545 | 11 (0-191) | 14 (0-147) | 0.81 |
| Baseline ESR, median (range) | 555 | 26 (1-110) | 28 (2-109) | 0.16 |
| Baseline DAS28, median (range) | 573 | 4.91 (0.77-8.23) | 5.28 (1.68-7.83) | 0.07 |
| HAQ, median (range) | 507 | 0.88 (0-2.75) | 1.13 (0-2.63) | 0.01 |
| DAS28 24 month AUC, median (range) | 450 | 83 (39-147) | 86 (33-146) | 0.94 |
| **Comorbidities** |  |  |  |  |
| Diabetes, n (%) | 103 | 2 (4) | 4 (14) | 0.19 |
| Hypertension, n (%) | 489 | 143 (38) | 48 (44) | 0.27 |
| Chronic heart disease, n (%) | 103 | 0 (0) | 1 (7) | 0.13 |
| TIA/stroke, n (%) | 103 | 1 (1) | 1 (7) | 0.25 |
| Coronary artery disease, n (%) | 103 | 4 (4) | 1 (7) | 0.53 |

While the presence of any CHIP was not significantly associated with RA disease activity, we hypothesized that specific gene mutations may be associated inflammation and enriched in patients with seropositive and seronegative disease subtypes. Patients with *DNMT3A* mutations had significantly higher ESR and disease-activity scores specifically in patients with seropositive, but not with seronegative, RA (Figure 5A-B); suggesting that *DNMT3A* may contribute to increased inflammatory activity in seropositive RA patients. The associations between *DNMT3A* and higher ESR and DAS28 were independent of age and sex (Supplementary Figure 13A-B) In contrast, *TET2* mutations were significantly more common in patients with seronegative RA both in a univariable (Figure 5C, Supplementary Figure 13C) and in a multivariable model that was adjusted for age, sex, and smoking (Figure 5D). A similar trend was observed across RA sub cohorts and when adjusting for ESR and CRP (Supplementary Figure 13D-F).

**Figure 5:**
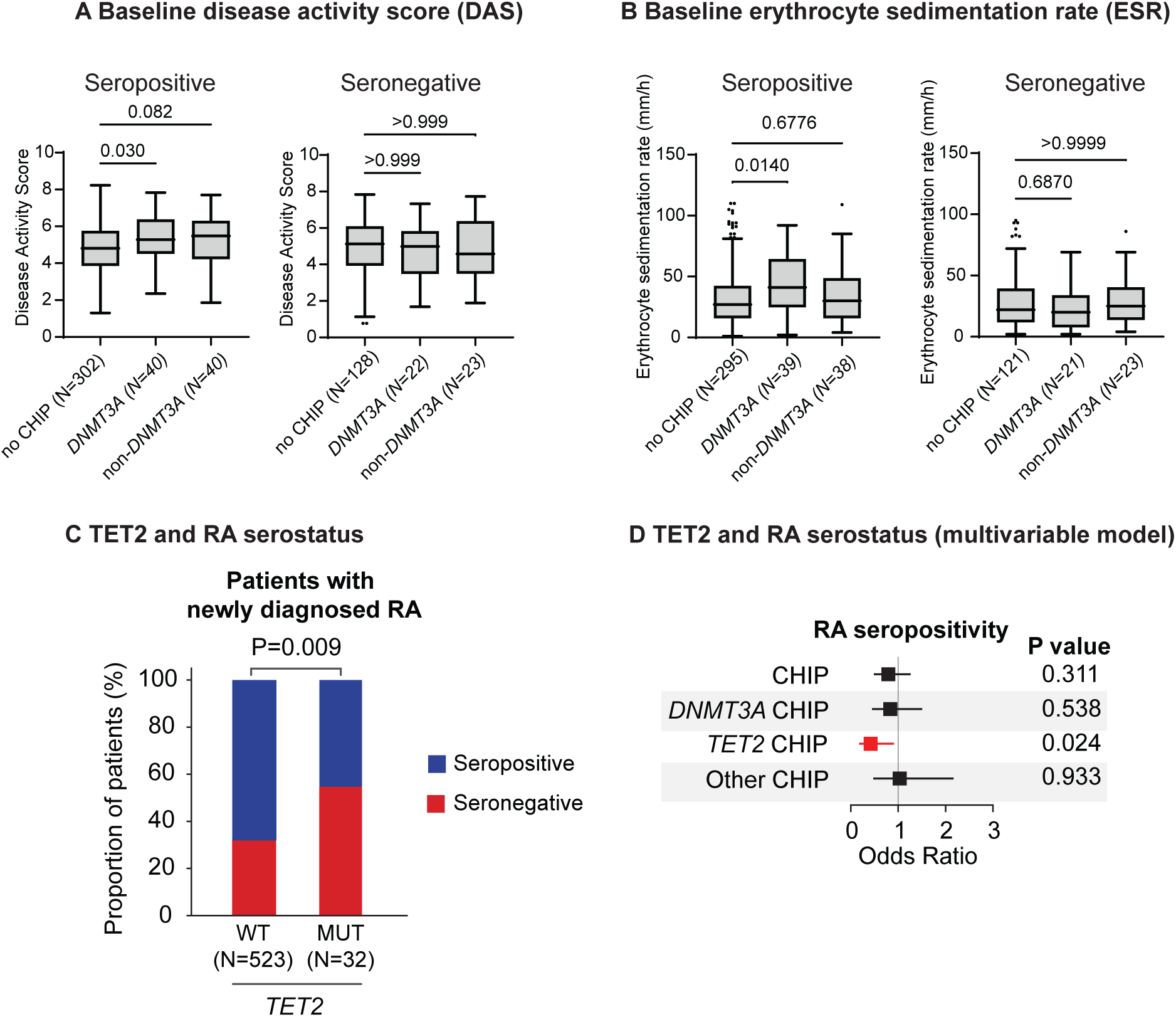
RA phenotypes at diagnosis associated with CHIP. (A) Disease activity scores by CHIP status in seropositive and seronegative RA. (B) Erythrocyte sedimentation rates by CHIP status and RA serotype. A-B P values calculated using Kruskal-Wallis test. (C) Proportion of seronegative RA patients by *TET2* CHIP status. The P value was calculated using Fisher’s exact test. (D) Multivariable model for CHIP subtypes and RA seropositivity adjusted for age, sex and smoking.

### CHIP and treatment outcomes in RA

The area under the curve for DAS28 during 24 months after RA diagnosis was similar between RA patients with and without CHIP (Table 1). Among 448 RA patients with long-term clinical follow-up, the median OS was 14 years. Patients with CHIP had similar OS compared with patients without CHIP (Figure 6A). However, we observed worse OS in seropositive RA patients with CHIP compared to seropositive patients without CHIP (Figure 6B). No difference in OS by CHIP status was observed in seronegative patients (Figure 6C). Furthermore, the cumulative incidence of cardiovascular events was similar in patients with and without CHIP (Figure 6D). No differences in OS or incidence of cardiovascular events by CHIP status were seen in multivariable models when adjusting for age, sex and smoking history (Figure 6E).

**Figure 6:**
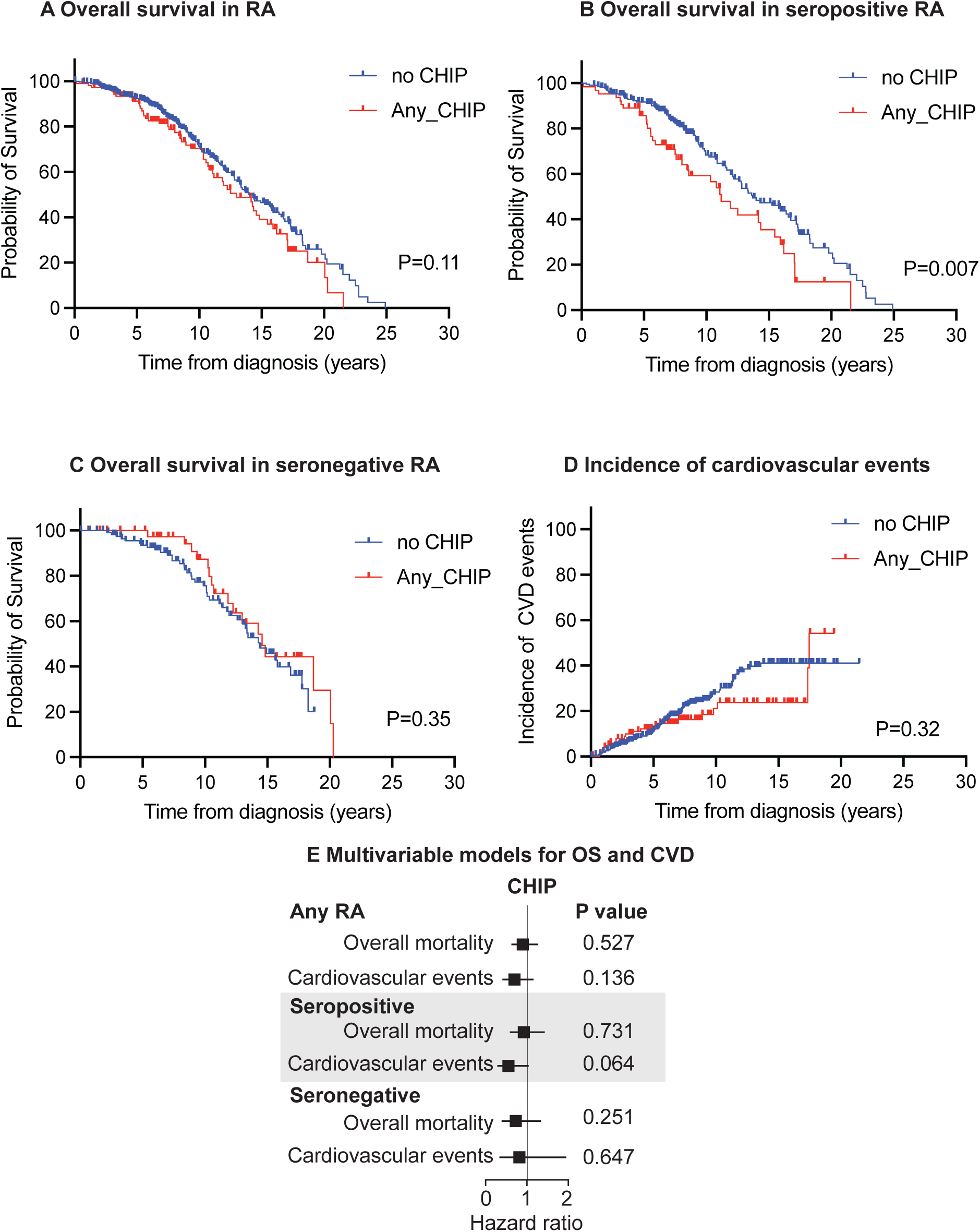
RA patient outcomes by CHIP status. (A) Kaplan Meier curve for overall survival in RA patients with and without CHIP. (B) Kaplan Meier curve for overall survival in seropositive RA patients with and without CHIP. (C) Kaplan Meier curve for overall survival in seronegative RA patients with and without CHIP. (D) Cumulative incidence of cardiovascular events in RA patients. (A-D) P-values calculated using Mantel-Cox test. (E) Cox proportional hazards models for overall mortality and cardiovascular events in RA patients. Adjusted for age, sex and smoking.

## Discussion

Our study adds to a growing body of evidence linking chronic inflammation with clonal HSC expansion in a context-specific manner^6^. In our data, CHIP was associated with prevalent RA in two population-level cohorts, consistent with smaller prior studies^25,26^. Understanding the interplay between age-related inflammatory diseases and clonal hematopoiesis may result in personalized therapeutic opportunities, as in the case of CANTOS trial, where carriers of *TET2* mutations were more likely to benefit from IL-1β inhibition for secondary CVD prevention^14^. IL-1β inhibition also belongs to the armamentarium of approved targeted therapies for RA^20^, and IL-1β blockage can suppress the expansion of *TET2* mutated HSCs in mice^12^. While we observed no differences in OS, CVD risk, or clinical remission rates by CHIP status in RA patients in multivariable models, the relative efficacy of IL-1β inhibition in RA subtypes by CHIP status could be tested in future clinical trials for personalized therapy.

Overall, CHIP was more common specifically in FinnGen participants with prevalent seronegative RA. Furthermore, in patients with newly diagnosed RA, *TET2* mutated CHIP was significantly enriched in patients with the seronegative disease subtype. Given the increased proinflammatory cytokine, particularly IL-1β, production in *TET2*-mutated monocytes/macrophages in murine models of gout^7^ and CVD^11,45^ as well as higher serum/plasma levels of proinflammatory cytokines in CHIP carriers^32^, our results collectively point towards a potential role for CHIP in promoting systemic inflammatory processes in seronegative RA. Production of IL-1 β from proinflammatory monocytes may act as key mediators of inflammation in RA pathogenesis^46^, and innate immunity is thought to play a more significant role in the pathogenesis of seronegative RA^47,48^. Intriguingly, a recent study identified proinflammatory M1 macrophages with upregulated IL1B gene expression in the synovia of ACPA-negative versus ACPA-positive RA patients^49^; the potential contribution of CHIP clones to this macrophage phenotype is an exciting area of future research. On the other hand, systematic inflammation in seronegative RA may selectively promote expansion of CHIP clones in the bone marrow and underlie the association between seronegative RA and CHIP.

Interestingly, *DNMT3A* mutated CHIP was more common in seropositive RA in the FinnGen cohort, and newly diagnosed seropositive RA patients with *DNMT3A* mutations had higher DAS disease severity score as well as higher ESR. A recent study by Wang and colleagues reported more severe arthritis phenotype and higher infiltration of *DNMT3A* mutated immune cells in the synovia of mice transplanted with HSC carrying the *DNMT3A* hotspot mutation and exposed to a collagen antibody-induced arthritis model^50^. On the other hand, our array-based CHIP analysis is unable to detect smaller clones and mutations in non-hotspot loci in the FinnGen cohort, which may bias the spectrum of CHIP mutations observed in RA subtypes. Despite opposing effects on DNA methylation^1^, both *Dnmt3a* and *Tet2* can promote proinflammatory macrophage phenotypes in atherosclerosis^51^. Further studies are warranted to understand the functional role and therapeutic implications of CHIP in distinct RA subtypes.

CHIP was associated with RA and distinct RA subtypes in all cohorts despite the heterogeneity in detecting CHIP across WES, SNP array, and targeted NGS methods, increasing the generalizability of our results. While seronegative RA is a heterogeneous disease entity^20^, the disease subtypes were carefully annotated by the treating rheumatologist in our RA patient cohorts. Subsequent studies with longitudinal blood DNA sampling pre- and post- RA diagnosis may help evaluate temporal clonal dynamics during disease development and upon anti-inflammatory therapy.

Together, our analyses using population-level and disease-specific cohorts point to a context-dependent association between CHIP and distinct RA subtypes. *DNMT3A* mutations were associated with a more severe clinical phenotype of seropositive RA, whereas *TET2*-mutated CHIP was enriched in the seronegative RA. Further studies are needed to delineate the clinical and functional relevance of CHIP detection in RA patients.

## Acknowledgements

This work was supported by the Sigrid Juselius Foundation (S.M, M.M), the Research Council of Finland (S.M, M.M), the Finnish Medical Foundation (M.M.), the European Research Council (Project: M-IMM 647355), Academy of Finland Heal-Art consortium (314442), Signe and Ane Gyllenberg Foundation (S.M), Helsinki Institute for Life Science Fellow Funding (S.M). For sequencing of RA patient samples, library preparation, sequencing and Dragen analysis were performed by FIMM Genomics NGS Sequencing unit at University of Helsinki supported by HiLIFE and Biocenter Finland. The center for Science Ltd. (CSC) is acknowledged for data storage and computational resources. FINRISK samples/data used for the research were obtained from THL Biobank (study number: BB2019_42). We thank all study participants for their generous participation at THL Biobank, and (a) specific cohort(s) as defined in IA. We acknowledge the Finnish Red Cross Blood Service for providing healthy control samples. We want to acknowledge the participants and investigators of the FinnGen study. The FinnGen project is funded by two grants from Business Finland (HUS 4685/31/2016 and UH 4386/31/2016) and the following industry partners: AbbVie Inc., AstraZeneca UK Ltd, Biogen MA Inc., Bristol Myers Squibb (and Celgene Corporation & Celgene International II Sàrl), Genentech Inc., Merck Sharp & Dohme LCC, Pfizer Inc., GlaxoSmithKline Intellectual Property Development Ltd., Sanofi US Services Inc., Maze Therapeutics Inc., Janssen Biotech Inc, Novartis AG, and Boehringer Ingelheim International GmbH. Following biobanks are acknowledged for delivering biobank samples to FinnGen: Auria Biobank (www.auria.fi/biopankki), THL Biobank (www.thl.fi/biobank), Helsinki Biobank (www.helsinginbiopankki.fi), Biobank Borealis of Northern Finland (https://www.ppshp.fi/Tutkimus-ja-opetus/Biopankki/Pages/Biobank-Borealis-briefly-in-English.aspx), Finnish Clinical Biobank Tampere (www.tays.fi/en-US/Research_and_development/Finnish_Clinical_Biobank_Tampere), Biobank of Eastern Finland (www.ita-suomenbiopankki.fi/en), Central Finland Biobank (www.ksshp.fi/fi-FI/Potilaalle/Biopankki), Finnish Red Cross Blood Service Biobank (www.veripalvelu.fi/verenluovutus/biopankkitoiminta), Terveystalo Biobank (www.terveystalo.com/fi/Yritystietoa/Terveystalo-Biopankki/Biopankki/) and Arctic Biobank (https://www.oulu.fi/en/university/faculties-and-units/faculty-medicine/northern-finland-birth-cohorts-and-arctic-biobank). All Finnish Biobanks are members of BBMRI.fi infrastructure (https://www.bbmri-eric.eu/national-nodes/finland/). Finnish Biobank Cooperative -FINBB (https://finbb.fi/) is the coordinator of BBMRI-ERIC operations in Finland. The Finnish biobank data can be accessed through the Fingenious^®^ services (https://site.fingenious.fi/en/) managed by FINBB.

## Authors Contributions

E.H., S.M., and M.M designed the study, analyzed data and wrote the manuscript. J.K., J.K., M.A., A.L, S.L., M.K., P.H., H.A., P.E., P.S., T.K., G.G., A.G. contributed to variant calling and mCA detection as well as curation of the genotype and phenotype data in the FinnGen and FINRISK studies. H.K., M.L-R., R.K., R.P., L.P., P.I., M.K., O.K-S., I.S., A.T.V, R.L., O.S., and S.R-D. collected and provided RA patient samples as well as clinical RA patient phenotypes. H.K. and S.R-D. also contributed to statistical analyses in the RA patient data. All authors reviewed the manuscript during its preparation and approved the submission.

