## Supplementary Figure for "Clonal hematopoiesis is associated with distinct rheumatoid arthritis phenotypes"

**Supplementary Figures**

Supplementary Figure 1: FINRISK cohort characteristics

Supplementary Figure 2: CHIP characteristics in FINRISK

Supplementary Figure 3: Lymphoid driver characteristics in FINRISK

Supplementary Figure 4: mCAs in FINRISK

Supplementary Figure 5: Associations between clonal hematopoiesis phenotypes

and selected phenotypes in FINRISK

Supplementary Figure 6: CH associated with RA in FINRISK

Supplementary Figure 7: CHIP hotspot variant calling in FinnGen

Supplementary Figure 8: Association between CHIP subtypes and RA subtypes in FinnGen

Supplementary Figure 9: Associations between mCA subtypes and RA subtypes

Supplementary Figure 10: CH associated with incident RA in FinnGen.

Supplementary Figure 11: CHIP characteristics in RA cohort

Supplementary Figure 12: Ca score by CHIP status in RA patients in ERA_CVD study (N=15)

Supplementary Figure 13: RA characteristics by CHIP status

**Supplementary Tables**

Supplementary Table 1: FINRISK with and without WES

Supplementary Table 2: CHIP candidate list

Supplementary Table 3: Lymphoid drivers candidate list. 3=Pathogenic, 2=Putative, 1=ALL

Supplementary Table 4: CHIP/Lymphoid driver variants detected in FINRISK

Supplementary Table 5: MCAs detected in FINRISK

Supplementary Table 6: ICD-code based phenotype definitions

Supplementary Table 7: SNP array derived-CHIP detection test in FINRISK

Supplementary Table 8: SNP array derived CHIP filtering results and criteria

Supplementary Table 9: RA patient cohort characteristics

Supplementary Table 10: Panel sequencing areas

Supplementary Table 11: CHIP variants detected in RA patient cohort

**Supplementary Figure 1: FINRISK cohort characteristics**

(A) Age and sex distribution in FINRISK. (B) RA cases in FINRISK. (C) Distribution of target coverage in FINRISK

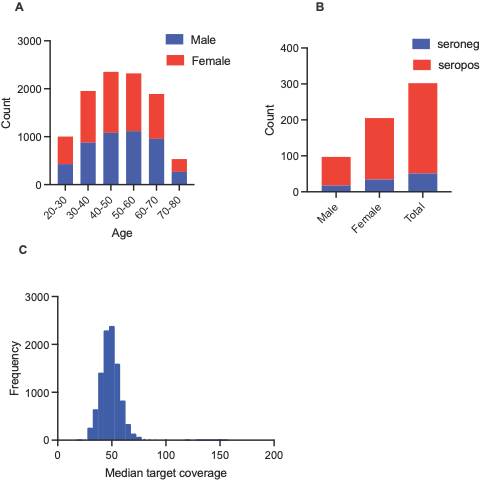

**Supplementary Figure 2: CHIP characteristics in FINRISK**

(A) Distribution of CHIP variant consequences. (B) Distribution of

nucleotide substitutions. (C) VAF distribution by gene. (D) VAF

distribution by variant consequence. (E) Female/Male ratio by gene.

(F) Empirical cumulative distribution of the most common gene

variants by age. (G) Number of participants with 1, 2 or 3 CHIP mutations.

(H) Comparison of coverages at CHIP gene exons in FINRISK and UK biobank (UKBB)

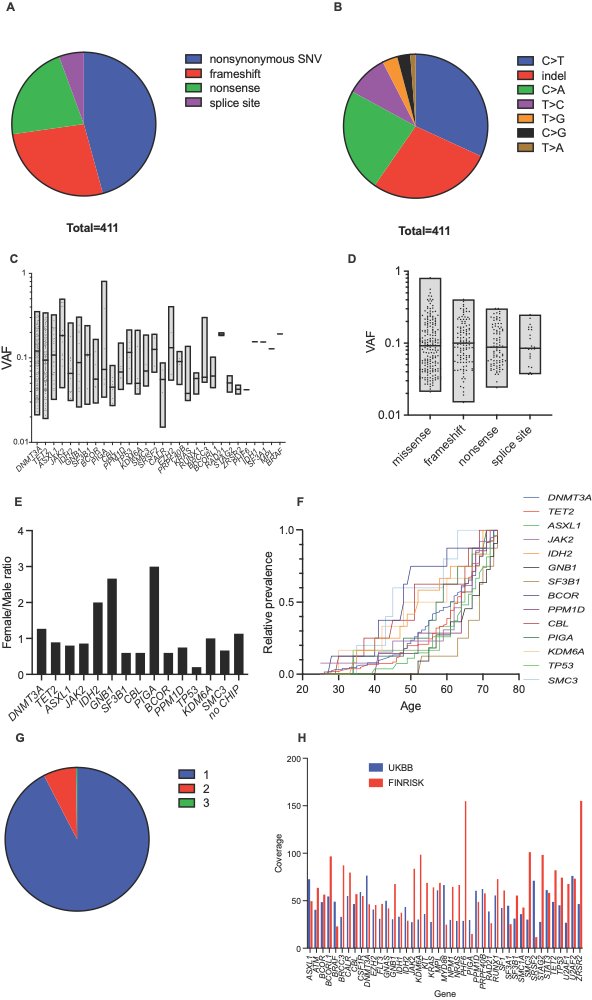
**Supplementary Figure 3: Lymphoid driver characteristics in FINRISK**

(A) Distribution of lymphoid driver variant consequences. (B) Distribution of

nucleotide substitutions. (C) VAF distributions by the most common lymphoid driver genes. (D) VAF distributions by variant consequence. (E) Number of participants with 1, 2 or more lymphoid driver variants. (F) CHIP vs lymphoid driver VAFs. C-F excluding prevalent hematologic malignancies.

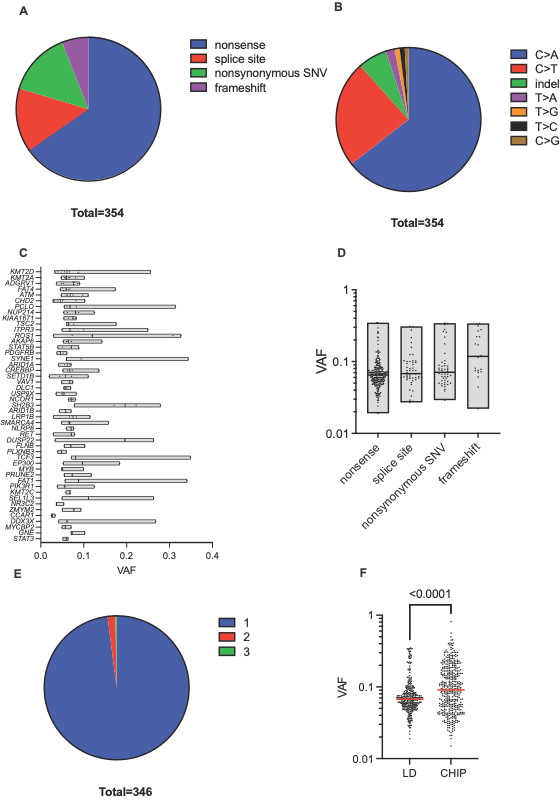

**Supplementary Figure 4: mCAs in FINRISK**

(A) Distribution of autosomal loss events by chromosome. (B) Distribution of autosomal CN-LOH events by chromosome. (C) Distribution of autosomal gain events by chromosome. (D) Distribution of estimated cell fractions by chromosome.

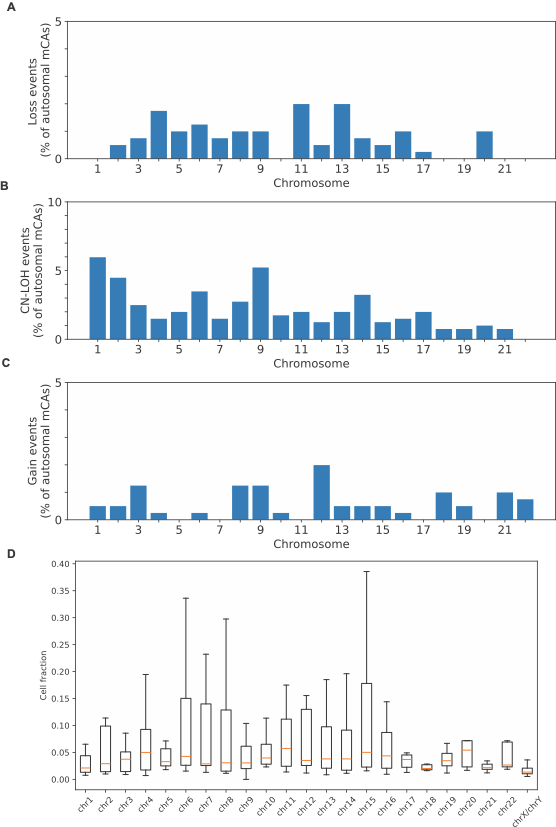

**Supplementary Figure 5: Associations between clonal hematopoiesis phenotypes and selected phenotypes in FINRISK**

(A) Associations of between CH and normalized blood counts and biochemical measurements. Calculations performed using a generalized linear model adjusting for age, sex, smoking history and principal components of ancestry, and excluding prevalent hematologic malignancies. (B) Associations between CH and smoking phenotypes. High, medium and low exposure defined as >=20, 5-20 and 0-5 pack years. Fitted using a logistic regression model excluding prevalent hematologic malignancies, not adjusting for covariates. (C) Association of CH and prevalent (i) myeloid and (ii) lymphoid malignancy diagnosis, fitted using a logistic regression model adjusting for age, sex, smoking and 10 principal components. (D) Association if CH and incident (i) myeloid and (ii) lymphoid malignancy diagnosis, using Cox-PR models. (E) CH and overall survival. D-E fitted using a Cox-PH model adjusted for age, sex, smoking history and PC:s of ancestry. A-B: *P<0.05, **P<0.01, ***P<0.00001. A: ALAT: alanine transaminase, CA: Serum calcium, CRP: C-reactive protein, GCT: Serum gamma-glutamyl transferase, GRA_N: Granulocytes (N), GRA_P: Granulocytes (%), HBA1C: Glycosylated hemoglobin, HBA1C_P: Glycosylated hemoglobin (%), HCT: Hematocrit level, HDL: Serum high-density lipoprotein, HGB: Hemoglobin, K: Urine potassium, KOL: Serum cholesterol, KREA: Serum creatinine, LDL: Serum low-density lipoprotein, LYM_N: Lymphocyte count, LYM_P: Lymphocytes (%), MCH: Mean cell hemoglobin, MCHC: Mean corpuscular hemoglobin, MCV: Mean erythrocyte volume, MPV: Platelet mean volume, PLT: Platelet count, RBC: Red blood cell count, RBV: Red blood cell size distribution width, TRIG: Serum triglyceride levels, U_ALB: Urine albumine, U_KREA: Urine creatinine, WBC: White blood cell count

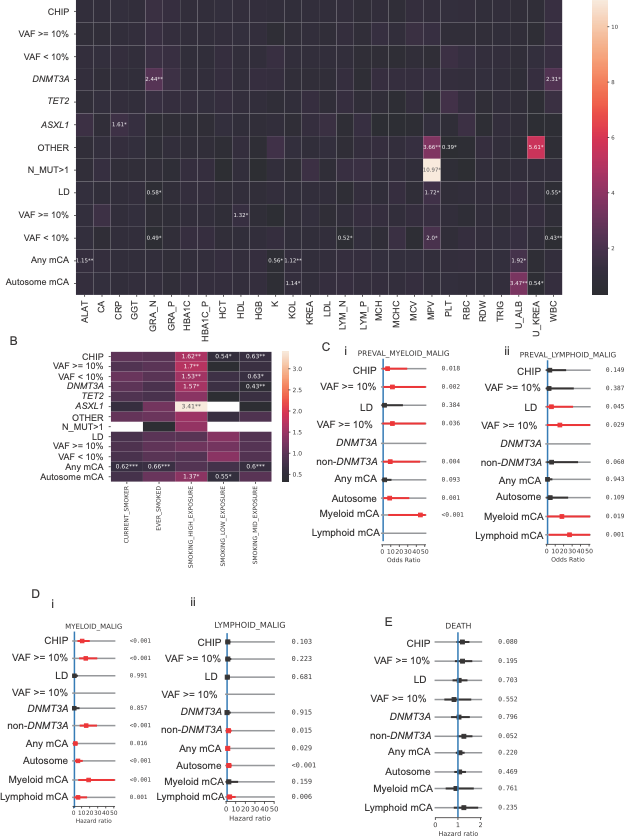

**Supplementary Figure 6: CH associated with RA in FINRISK**

(A) CH associated with seropositive RA (B) CH associated with seronegative RA (C) CH associated with incident RA (Cox-PH model) (D) CH associated with questionnaire-based RA

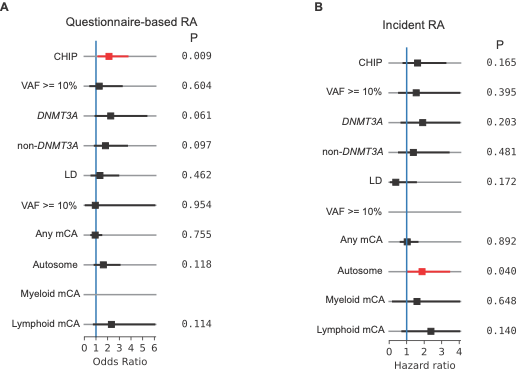

**Supplementary Figure 7: CHIP hotspot variant calling in FinnGen**

(A) Method for calling and filtering CHIP hotspot loci in FinnGen. (B) Age association by CHIP gene in FinnGen. (C) Association with prevalent hematologic malignancies. (D) Association with incident hematologic malignancies. (E) Association with C-reactive protein levels.

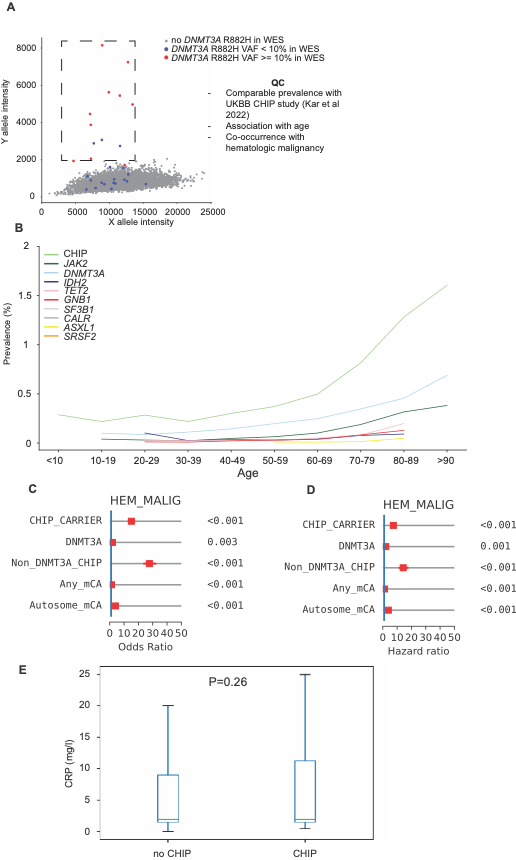

**Supplementary Figure 8: Association between CHIP subtypes and RA subtypes in FinnGen**

(A) RA case counts in FinnGen. (B) Association of CHIP subtypes with prevalent RA. (C) Association of CHIP subtypes with prevalent seropositive RA. (D) Association of CHIP subtypes with prevalent seronegative RA. Models adjusted for age, sex, smoking and 10 principal components of ancestry, excluding prevalent hematologic malignancies.

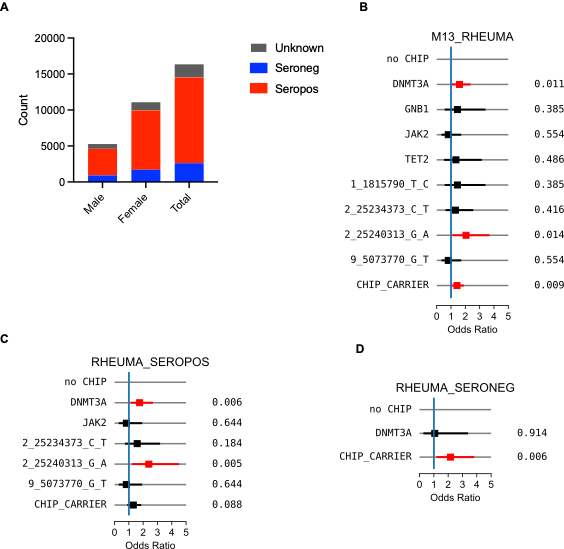

**Supplementary Figure 9: Associations between mCAs and RA subtypes**

(A) Association between mCA subtypes and prevalent RA. (B) Association between

mCA subtypes and prevalent seropositive RA. (C) Association between mCA subtypes and seronegative RA.

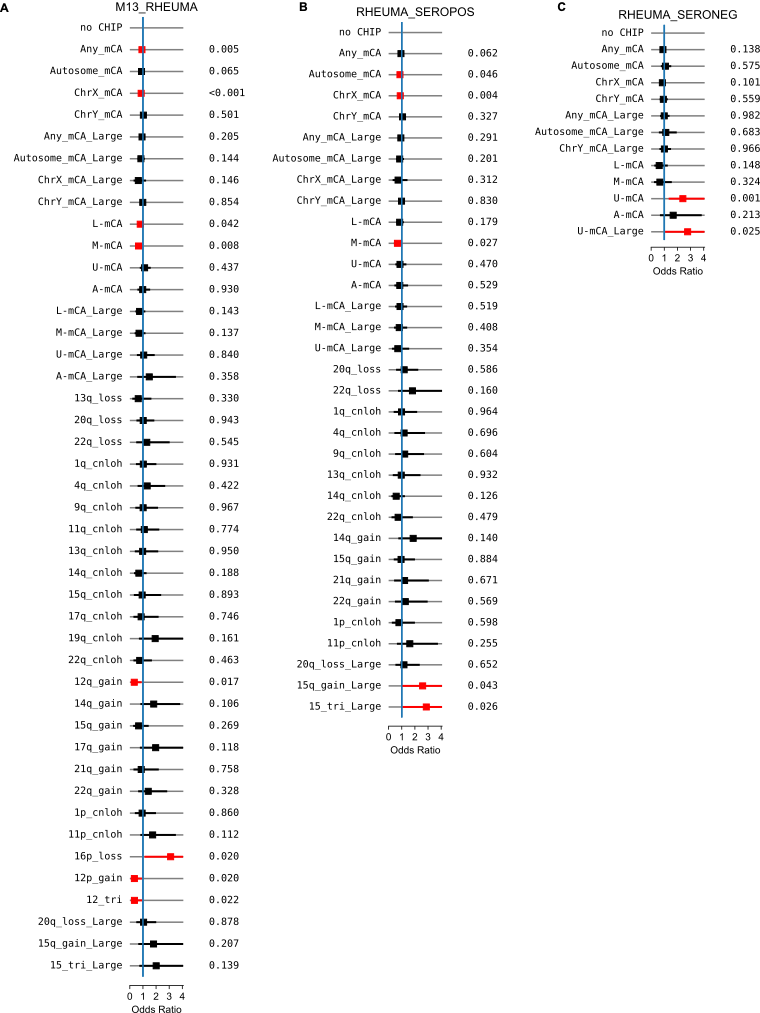

**Supplementary Figure 10: CH associated with incident RA in FinnGen.**

Cox-PH models adjusted for age, sex, smoking, 10 principal components, and censoring hematologic malignancies

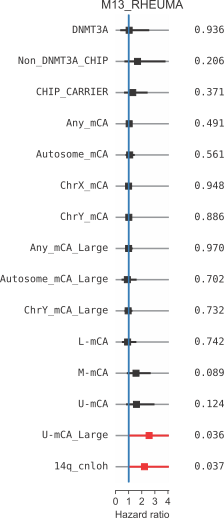

**Supplementary Figure 11: CHIP characteristics in RA cohort**

(A) Comutation plot for RA patients. (B) Number of participants with 1, 2,

or more CHIP variants. (C) CHIP status in RA vs healthy controls. Model adjusted for age and sex (D) CHIP prevalence by age in RA patients and healthy controls

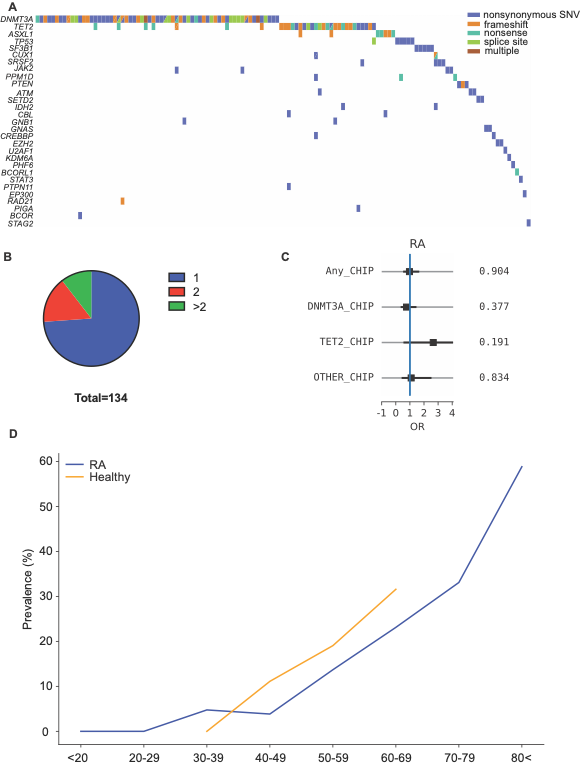

**Supplementary Figure 12: Ca score by CHIP status in RA patients in ERA_CVD study (N=15)**

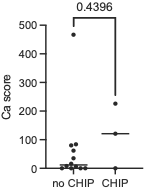

**Supplementary Figure 13: RA characteristics by CHIP status**

(A) Multivariable association between CHIP subtypes and ESR in seropositive RA patients. (B) Multivariable association between CHIP subtypes and ESR in seropositive RA patients. A-B fitted using a least squares model adjusted for age and sex. (C) Univariable associations between CHIP subtypes and RA serostatus. P-values calculated using Fisher’s exact test (D) Multivariable model for RA serostatus by CHIP status adjusted for age, sex, smoking, ESR and CRP. (E) Multivariable model for RA serostatus in Umea1 cohort. (F) Multivariable model for RA serostatus in Umea2 cohort. D-F adjusted for age, sex, smoking

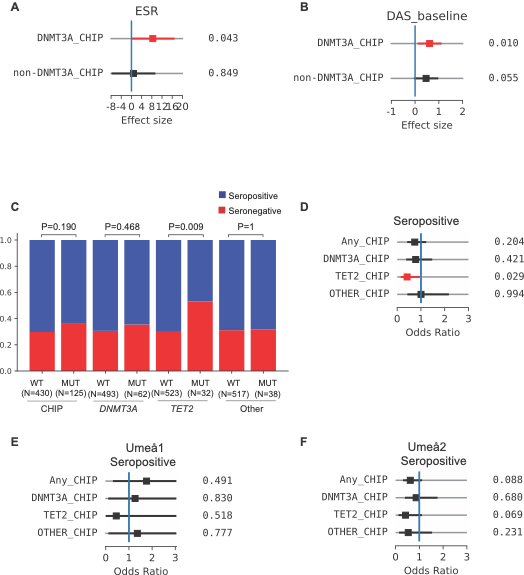
