## Supplementary Methods for "Clonal hematopoiesis is associated with distinct rheumatoid arthritis phenotypes"

*Variant calling and filtering in FINRISK*

For variant calling, we used DRAGEN 3.8 with parameters “–vc-systematic-noise-method aggregate and --vc-systematic-noise-use-germline-tag true” to generate systematic noise representing the panel of normals. For variant calling in tumor only mode, we used parameters –enable-map-align true, --enable-map-align-output true, --enable-duplicate-marking true, --pair-by-name true, --tumor-bam-input <bamfile> --enable-variant-caller true, -r <hg38-reference.fa> –auto-detect-sample-sex true, --enable-vcf-compression true, --vc-orientation-bias-filter-artifacts C/T, --vc-systematic-noise <systematic-noise-file> --vc-forcegt-vcf <cosmicmutations.vcf> --dbsnp <dbsnp-20180418> --soft-read-trimmers quality, adapter, --trim-min-quality 3, --trim-min-length 20, --trim-adapter-read1 <bbduk_adapters.fa>, --trim-adapter-read2 <bbduk_adapters.fa>, --enable-sv true, --sv-exome true, --enable-cnv true, --cnv-population-b-allele-vcf <gnomad_v3_popaf.0.10.sort.vcf.gz>, --cnv-use-somatic-vc-vaf=false, --cnv-target-bed <VCRome_2_1_hg38_capture_targets_padding50.bed>, --cnv-normals-list <cnv_normals_list.txt>

To detect CHIP variants, we used a candidate list consisting of 96 genes (Supplementary table 2). To filter benign and likely germline variation, we used a maximum population allele frequency threshold of 0.1% in WES of any population in gnomAD^1^ (version 3.1.2), excluding known CHIP hotspots *JAK2* V617F and *DNMT3A* R882. In addition, we required variants to be labeled “PASS” by DRAGEN, have a somatic quality score 3 or higher, a GermlineQuality score of less than 10 and have at least 10 reads aligning to the variant loci, out of which at least 2 are variant reads. At least one read on both strands supporting both alleles was required. Furthermore, splicing variants occurring further than 2 bp from the intron-exon boundary were excluded, as well as frameshift variants seen in more than 0.05% of samples. For frameshift variants occurring within 5 bp of each other, only the variant with higher variant allele frequency (VAF) was included for downstream analysis. No VAF thresholds were used to filter variants. Finally, we manually curated detected variants using IGV.

For lymphoid driver variants, we generated a list of 493 candidate genes described in supplementary table 3, containing the genes and candidate loci originally described by Niroula et al as lymphoid CHIP appended with candidate loci of known acute lymphoblastic leukemia (ALL) driver genes^2^. Variants identified in these genes were filtered as above. In addition, for lymphoid driver variants, we required the VAFs to be between 2% and 35% and for missense lymphoid driver variants, we required the variant to be reported at least three times in COSMIC or at least once in the context of L-CHIP^3^ or ALL^2^.

*FinnGen cohort*

FinnGen samples are collected from six regional hospital biobanks, a national level biobank, a private sector biobank and the Blood Service biobank. The majority of FinnGen samples have been genotyped using FinnGen’s custom ThermoFisher Axiom array, containing 736 145 probes for 655 973 loci. In addition, a subset of samples was genotyped using legacy chips from old studies. To further enhance the genetic information available, further SNPs were imputed using the SISu v.4.0 imputation panel.

*CHIP calling in FinnGen*

For candidate SNP loci in CHIP genes, filtering criteria were defined as follows: less than 10-fold prevalence of variants relative to UKBB data, positive univariate (P<0.1) association with age and positive univariate association (P<0.05) with either myeloid, lymphoid or other hematologic malignancy event in the registry (Supplementary Table 6). If the relative prevalence was less than 2-fold, we required one of the associations with age or hematologic malignancy.

*RA patient cohorts*

We selected two distinct RA patient cohorts from the Biobank at the Rheumatology department in Umeå, Sweden. Cohort 1 consisted of 150 RA patients that were selected in a case-control manner for CVD^4^. Cohort 2 consisted of 300 early RA patients, half of which had seronegative (ACPA and RF negative) and half seropositive (ACPA and/or RF positive) RA subtypes^5^. In addition, we included samples from 18 and 60 RA patients from Finnish FIN-RACo^6^ and NEO-RACo^7^ clinical trials, respectively, as well as from a prospective cohort of 45 patients with blood sample collection at RA diagnosis (FosfoRA). Furthermore, we included 59 samples with CHIP information available, from a previously published cohort study on CHIP and RA by Savola et al^8^. In addition, we sequenced 163 healthy controls. All participants provided informed consent to participate in the study.

*Targeted next-generation sequencing panel to detect CHIP*

40-50 ng of gDNA was processed according to Twist Library Preparation EF 2.0 with Enzymatic Fragmentation DOC-001239 REV 1.0 and Twist Target Enrichment Protocol DOC-001085 REV 2.0 manual (Twist Bioscience, San Francisco, CA, USA) with following modifications. IDT xGEN unique dual index (UDI) with UMI (unique molecular identifier adapters were used for ligation (Integrated DNA Technologies, Coralville, IA, USA). Library quantification and quality check was performed using LabChip GX Touch HT High Sensitivity assay (PerkinElmer, Waltham, MA, USA) and Qubit Broad Range DNA Assay (Thermo Fisher Scientific, Waltham, MA, USA). Libraries were pooled to 14-16-plex reactions. The exome enrichment was performed using Twist custom panel probes (244KB). The captured library pools were quantified for sequencing using QuantStudio5 Collibri Library Quantification kit (Thermo Fisher Scientific, Waltham, MA, USA) and LabChip GX Touch HT High Sensitivity assay (PerkinElmer, Waltham, MA, USA).

Sequencing was performed with Illumina NovaSeq 6000 system (Illumina, San Diego, CA, USA) and v1.5 chemistry. The median target sequencing coverage was 1700x across samples.

We used Illumina DRAGEN 3.8 in tumor-only mode for variant calling and ANNOVAR for variant annotation. The DRAGEN parameters we used were –ref-dir <refdir>, --output-file-prefix <sampleID>, --tumor-fastq1 <sampleID>.R1.fastq.gz, --tumor-fastq2 <sampleID>.R3.fastq.gz –RGID-tumor <rgid> --RGSM-tumor <rgsm> --enable-sort true, --enable-map-align-output true, --enable-duplicate-marking false, --enable-variant-caller true, --vc-enable-joint-detection true, --vc-systematic-noise <WGS_hg38_v1.0_systematic_noise.bed>, --vc-enable-umi-solid true, --vc-snp-error-cal-bed <hg38_probe_area.bed> , --high-coverage-support-mode true, --enable-sv true, --sv-exome true, --sv-systematic-noise <WGS_v1.0.0_hg38_sv_systematic_noise.bedpe.gz>, --umi-enable true, --umi-library-type random-simplex, --umi-min-supporting-reads 1, --umi-source fastq, --umi-fastq <sampleID>. R2.fastq.gz , --umi-metrics-interval-file < hg38_probe_area.bed>, --umi-correction-scheme random, --qc-coverage-reports-1 full_res, --qc-coverage-region-1 <hg38_probe_area.bed>

Only variants with “PASS” label from DRAGEN were included for subsequent filtering. We excluded variants with gnomAD maximum population allele frequency of over 0.1% or occurring in over 1% of our samples. In addition, we excluded variants with less than 100 total reads, or 2 variant allele reads. We excluded variants with a VAF of over 40%. Furthermore, we required every variant to have at least one read in each direction supporting each allele. Finally, we included only variants with a variant allele frequency of at least 2% as CHIP, although variants with VAFs as low as 0.5% were included for some exploratory analyses.
